# Exploring stakeholder perspectives on the content, delivery, and contextual delivery of shared decision-making for people living with subacromial pain syndrome: a multimethod study with future workshops and interviews

**DOI:** 10.64898/2026.07.02.26356879

**Authors:** Kristian Damgaard Lyng, Simon Kristoffer Johansen, Nadine E Foster, Jens Lykkegaard Olesen, Janus Laust Thomsen, Jens Søndergaard, Michael Skovdal Rathleff

## Abstract

**Background:** Shared decision-making (SDM) is a key component in patient-centered care for people consulting health care due to chronic musculoskeletal pain, including subacromial pain syndrome (SAPS). Limited research has explored how patients, relatives, and healthcare professionals perceive the content and delivery of SDM for managing SAPS in primary care. Thus, this study aims to explore stakeholder perspectives on the content, delivery, and contextual requirements for a context-specific SDM intervention for SAPS, and to identify shared challenges and co-develop ideas to inform intervention development.

**Methods:** We conducted three separate future workshops (patients/relatives, physiotherapists/chiropractors, and general practitioners), each consisting of structured critique, fantasy, and implementation phases. A rapid preliminary analysis of workshop data was followed by semi-structured stakeholder interviews to validate, challenge, or elaborate the findings. All data were analysed thematically using an iterative, reflexive approach.

**Results:** Twenty-eight participants took part across three workshops: patients/relatives (n = 10), physiotherapists/chiropractors (n = 12), and general practitioners (n = 6). Six additional stakeholders provided inputs via subsequent interviews (three physiotherapists, one patient, one relative and one GP). Thematic analysis identified 20 themes and 59 sub-themes, which were refined into two overarching categories: (1) shared barriers to SDM in SAPS care, including diagnostic uncertainty, fragmented clinical care pathways, time constraints, and decision fatigue; and (2) stakeholder visions for future SDM interventions, emphasising continuity, tailored communication tools, and supportive digital ecosystems.

**Conclusion:** Based on stakeholder input, SDM in SAPS care may consider integrating dynamic, integrated systems that account for diagnostic ambiguity, contextual constraints, and varying patient capacities. These findings provide an actionable foundation for co-developing and piloting a context specific SDM intervention for primary care.

## Introduction

Shared decisionLJmaking (SDM) is an approach in which clinicians and patients work together to make healthcare decisions (Elwyn et al., 2025). Clinicians present the best available evidence, and patients are supported in weighing the options considering their values, preferences, and circumstances, with the goal of reaching an informed and mutually agreed choice (Montori et al., 2023). SDM is particularly relevant in preference-sensitive conditions, where multiple reasonable management options exist and no single option is appropriate for every patient (Elwyn et al., 2009; Franco et al., 2020). Subacromial pain syndrome (SAPS) is the most common cause of shoulder pain, affecting 36–48% of patients in primary and secondary care of those presenting with shoulder pain (Diercks et al., 2014; Juel and Natvig, 2014; Lucas et al., 2022; Windt et al., 1995). Although exercise remains the recommended first-line treatment, uncertainty remains regarding the optimal delivery, intensity, and sequencing of non-surgical management, making treatment decisions highly dependent on individual patient goals, preferences, and circumstances (Powell et al., 2024, 2022).

Although SDM is generally supported by systematic reviews and recommended by healthcare policy makers (Bruch et al., 2024; Stacey et al., 2024), there is currently no SDM - specific interventions for SAPS, and little evidence to guide how SDM should be integrated into routine SAPS care. Our previous work has identified SDM as a research priority for SAPS, demonstrated that patients experience substantial decisional needs when navigating management options, and shown that a context-specific decision aid is both feasible and acceptable in primary care (Samantha C. Bengtsen et al., 2025; Samantha Charmaine Bengtsen et al., 2025a; Lyng et al., 2025, 2026). However, a decision aid alone is unlikely to support meaningful implementation of SDM in clinical practice. Our realist review further highlighted that successful SDM depends on the interaction between intervention components, stakeholder behaviours, and the context in which care is delivered (*unpublished data*). Together, these findings suggest that important knowledge gaps remain regarding how SDM should be designed and delivered for patients with SAPS.

Understanding how patients, relatives, and healthcare professionals perceive the content, delivery, and implementation of SDM is therefore a necessary step before refining and evaluating a context-specific intervention. The aim of this study was to explore stakeholder perspectives on the content and delivery of SDM for patients living with SAPS to inform the refinement of a context-specific SDM intervention specifically designed for primary care in Denmark.

## Methods

### Study design and rationale

This study employed a multi-stage qualitative design involving three workshops, conducted separately with general practitioners (GPs), physiotherapists and chiropractors, and patients with SAPS, followed by semi-structured stakeholder interviews (**See figure 1**). The future workshop method, originally developed by Jungk and later refined by Vidal, was used to identify current challenges, generate future visions, and co-develop potential solutions to support SDM in SAPS care (Jungk and Muellert, 1987; Vidal, 2005). To strengthen the credibility and transferability of the findings, additional stakeholder interviews were conducted with individuals who had not participated in the workshops. These interviews served as an external stakeholder validation, allowing emerging themes and proposed intervention ideas to be validated, elaborated upon, or challenged from independent perspectives and across broader clinical contexts (Birt et al., 2016). The study was guided by the UK Medical Research Council (MRC) framework for developing and evaluating complex interventions, which emphasises iterative intervention development informed by stakeholder engagement and contextual understanding (Skivington et al., 2021). Reporting of the study is inspired by the COnsolidated criteria for REporting Qualitative research (COREQ) checklist (Tong et al., 2007).

**Figure 1.**
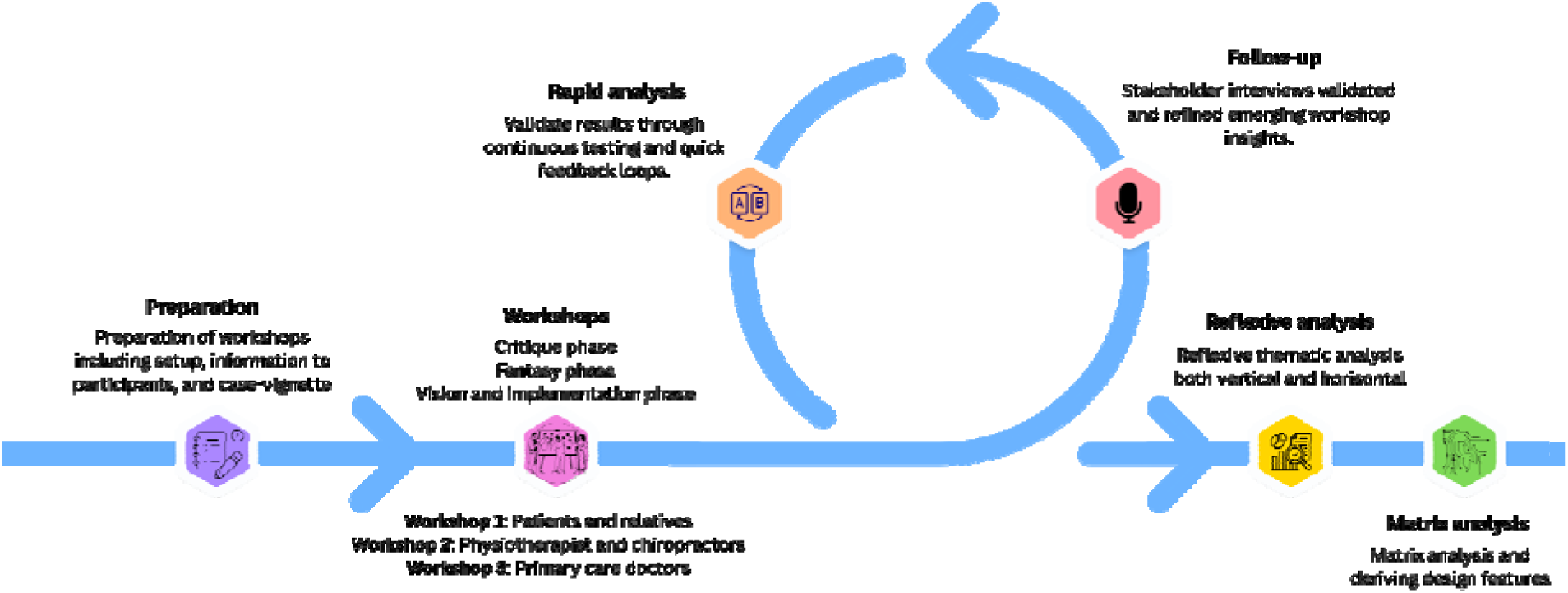
Study process outlining all steps of the study.

### Context and setting

The study was conducted primarily within the Region of Northern Jutland, Denmark, but included participants from across the country to enhance the relevance and transferability of the findings. The Danish healthcare system is publicly funded and organised around primary care, where general practitioners (GPs) serve as gatekeepers to publicly funded specialist services, including hospital-based care and referrals to secondary care. Private physiotherapists and chiropractors also play a central role in the assessment and management of musculoskeletal conditions and are commonly involved in the treatment of patients with SAPS. Healthcare professionals were recruited primarily from the Region of Northern Jutland, reflecting the regional organisation of primary care services, whereas patients with SAPS were recruited nationally to capture a broader range of lived experiences.

### Participant recruitment and sampling strategy

We purposively recruited four stakeholder groups including 1) patients with SAPS, 2) family members or informal carers, 3) physiotherapists and chiropractors, and 4) GPs (See **Table 1**). These groups were selected to capture a range of perspectives relevant to the development and implementation of SDM interventions for SAPS. Patients were recruited nationally through social media, patient associations, and targeted outreach. GPs were recruited within the Region of Northern Jutland via professional clinician networks, including the Center for General Practice at Aalborg University and Nord-KAP (the regional quality unit for general practice), email invitations, and social media. Physiotherapists and chiropractors were similarly recruited through professional and local networks. All participants were aged 18 years or older. Healthcare practitioners were required to have at least two years of experience managing patients with SAPS. Patients needed to have current or previous experience with non-traumatic shoulder pain described as SAPS and current or previous healthcare consultations for their pain. Carers or family members were eligible if they had supported someone with SAPS in a relational or caregiving capacity. Potential participants who responded positively to our inclusion efforts were contacted via telephone at the time of their choosing, screened, and provided with verbal and written information in accordance with the stipulations in the European General Data Protection Regulations.

**Table 1.** Inclusion Criteria.

| Group | Criteria |
| --- | --- |
| Patients currently living with or have had a lived experience with SAPS | + 18 years of age<br>Diagnosed with non-traumatic shoulder pain, termed SAPS or similar |
| Family or carer | • + 18 years of age<br>• Relative or carer of a person currently living with or have had a lived experience with SAPS (e.g., family member, spouse). |
| Health-care practitioners | • + 18 years of age<br>• Registered health-care practitioner (e.g., general practitioner, physiotherapist, or chiropractor) |

### Procedure

Three workshops were conducted, one workshop with patients with SAPS and relatives, one with physiotherapists and chiropractors, and lastly, one with GPs. All workshops consisted of three phases: critique phase, fantasy phase, and a vision and implementation phase as seen in similar studies in musculoskeletal pain (Johansen et al., 2023, 2022; Larsen et al., 2024; RB et al., 2025). In addition, each workshop was preceded by a preparatory phase, during which participants were introduced to the purpose and structure of the workshop. Following the workshops, a brief follow-up phase was conducted, allowing participants to reflect on the experience, clarify key outputs, and provide additional feedback to inform subsequent analysis and interpretation. Following completion of the workshops, an independent stakeholder validation phase was conducted through semi-structured interviews with stakeholders who had not participated in the workshops. These interviews were used to validate, elaborate on, or challenge the emerging themes and proposed intervention concepts, thereby extending the findings beyond the original workshop participants. The individual stages of the study are described below. Two pilot workshops were conducted prior to the preparatory phase to familiarise the facilitator (KDL) with the format. All participants received I) a short survey to capture key demographic data and II) a short protocol on the future workshops that aims to familiarise participants with the approach and the phases within. All participants were informed prior to the future workshops that the workshop consisted of three phases and that the workshops had a duration of approximately 3 hours including breaks (See **Appendix 1** for example of time schedule). To support the transition between phases and inspire discussion through dialogue, a fictional case vignette was developed and introduced to illustrate a realistic scenario in which a SDM intervention might be relevant (**Appendix 2**). The vignette was informed by our previous qualitative work but does not represent any individual participant or real patient. Inspiration cards were used to stimulate creativity and support dialogue during the future workshops (See **Figure 2**). Each card featured an evocative image designed to represent themes such as uncertainty, communication, and collaboration, reflecting early insights from our previous work on SAPS and SDM.

**Figure 2.**
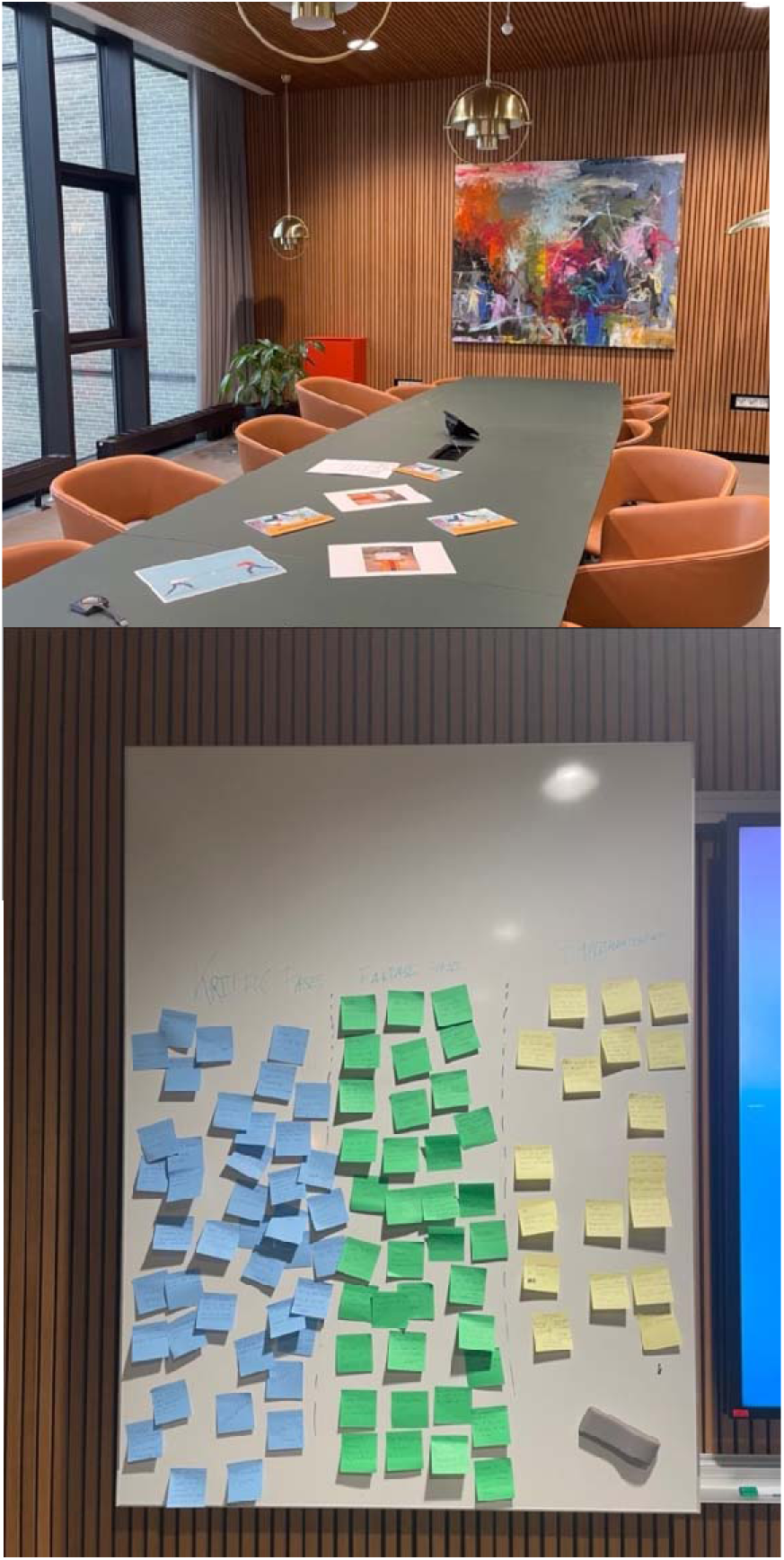
Setup for the workshops and example of data collection.

### Workshop phases

The future workshops were conducted using a structured four-phase process consisting of preparation, critique, fantasy, and vision/implementation, followed by an independent stakeholder validation phase. The objectives, activities, and outputs of each phase are summarised in **Table X**.

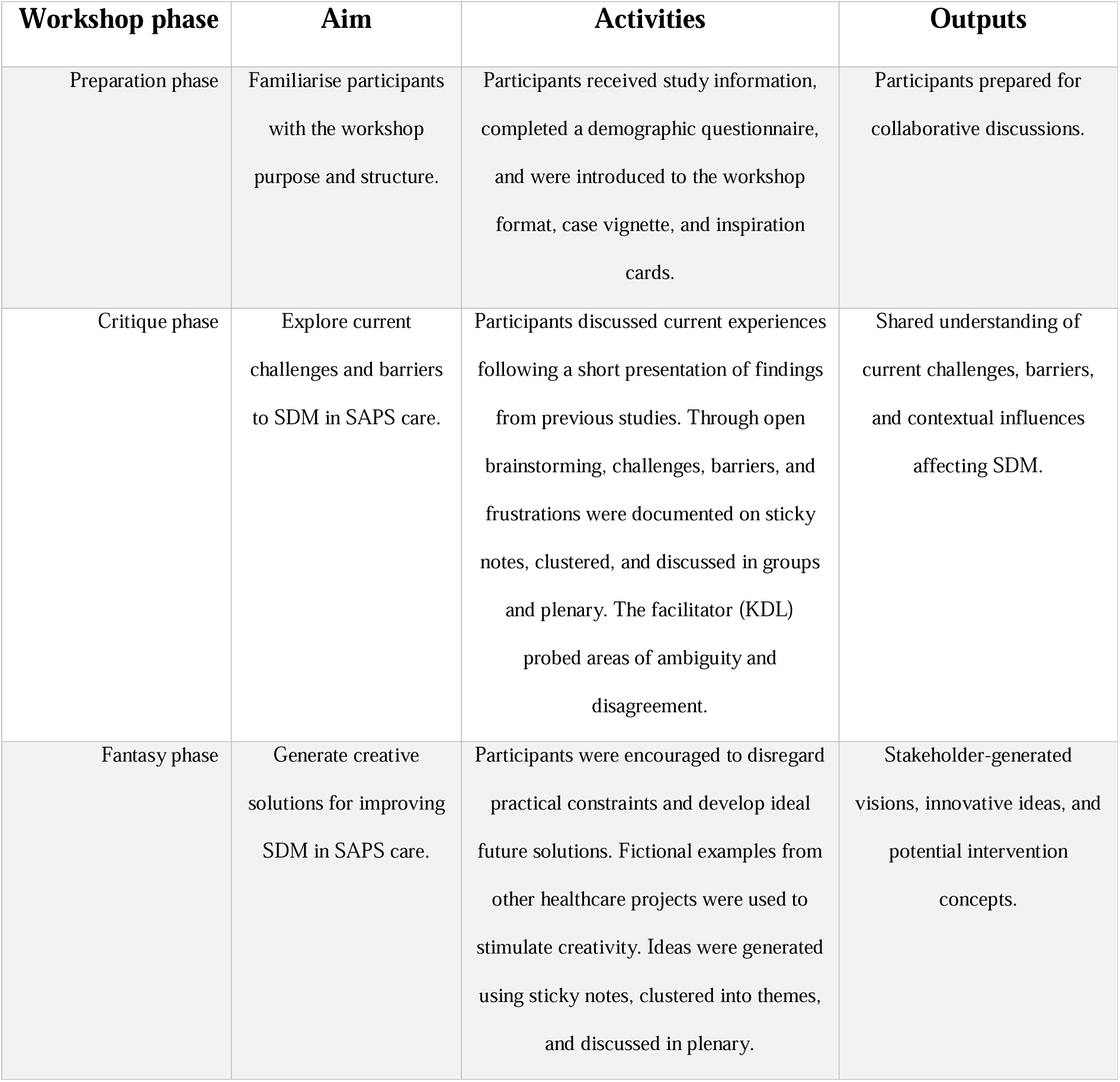
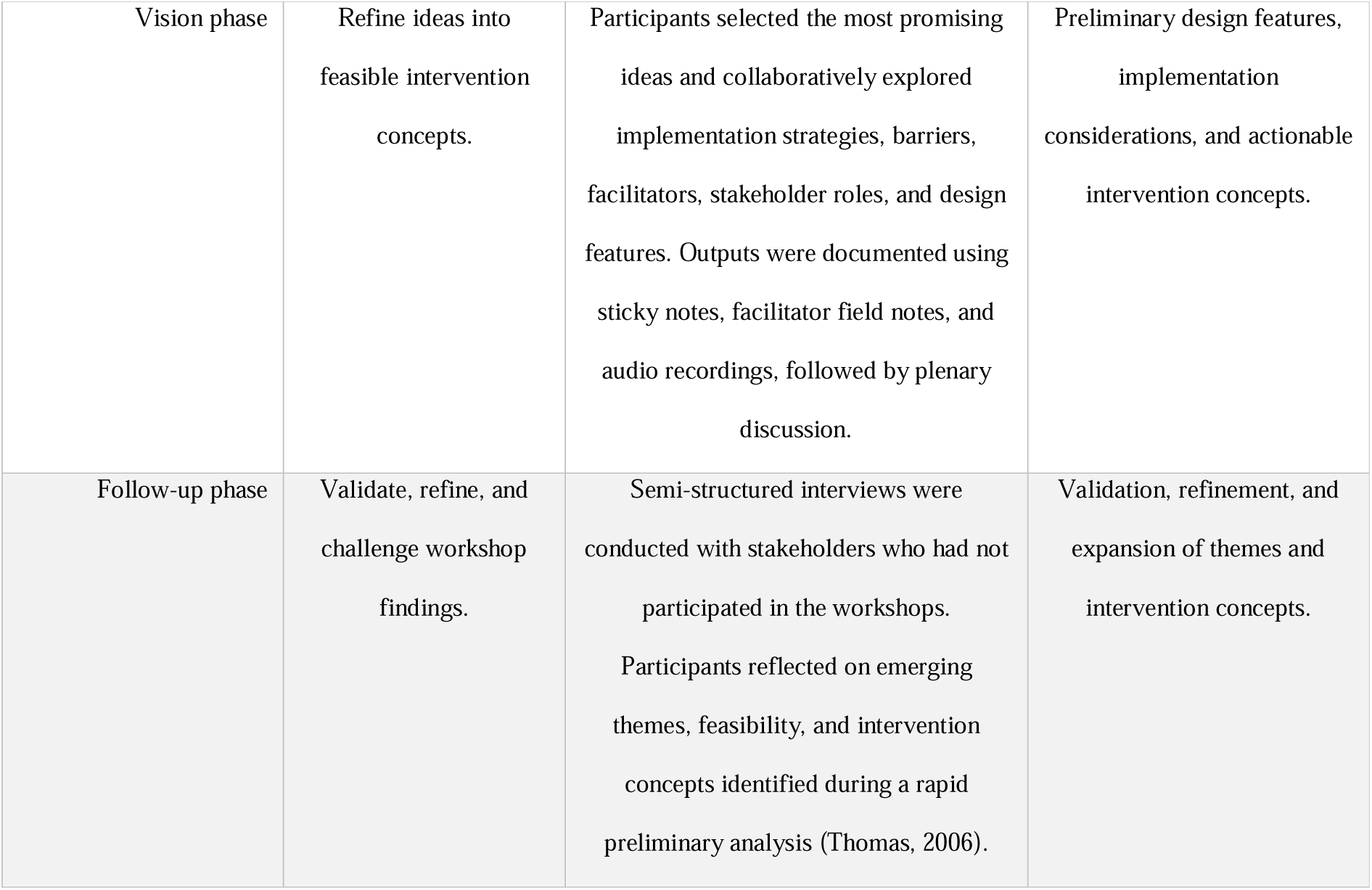

### Data management

Demographic data were collected using the secure web-based application REDCap™ (Research Electronic Data Capture), hosted by Aalborg University, Denmark (Harris et al., 2019, 2009). All workshops were audio recorded and transcribed using Whisper Transcription (version 1.1) and subsequently reviewed and corrected manually by the KDL to ensure accuracy. All transcripts were anonymised, with identifiable information removed during transcription. Anonymised transcripts, demographic data, and other research materials (e.g., facilitator notes, post-it outputs) were stored securely through Aalborg University in compliance with institutional data protection policies and the European General Data Protection Regulation (GDPR). Only authorised members of the research team had access to the raw data. Data will be retained for a period of five years following study completion, in accordance with Aalborg University’s research data management policies and applicable legal requirements. No personally identifiable data will be used in publications or presentations.

### Data analysis

All workshop and interview transcripts were analyzed using reflexive thematic analysis as described by Braun and Clarke (Braun and Clarke, 2006). Thematic text analysis consists of six phases: 1) familiarisation, 2) generation of initial codes, 3) searching for themes, 4) reviewing themes, 5) defining themes and, 6) writing up the findings. Both vertical patterns (within individual workshops) and horizontal patterns (across workshops and stakeholder interviews) were explored to capture group-specific experiences and shared dynamics related to SDM in SAPS (Clarke et al., 2019). Analysis proceeded iteratively and inductively, beginning with repeated readings of the data, followed by open coding of meaningful segments. Codes were then collated into preliminary categories, which were refined into overarching themes capturing challenges, visions, solutions, and contextual influences on SDM in SAPS care from each workshop. Outputs from sticky note clustering, facilitator field notes, and workshop summaries were integrated with the transcripts to support triangulation across data sources. Findings from the stakeholder interviews were analyzed in the same manner and used to validate, elaborate on, or challenge themes emerging from the workshops. Following thematic analysis, a matrix analysis was conducted to systematically compare themes across the three workshops and stakeholder interviews. This step enabled identification of convergence, divergence, and tensions across actors, domains, and levels of the healthcare system, and supported synthesis of the findings into a conceptual model capturing key relationships and pressure points in SDM for SAPS. Building on this model, and the stakeholder-generated visions articulated during the workshops, empirically grounded design principles and concrete design features for a future SDM intervention were derived.

### Ethical considerations

The separate workshops were chosen to avoid potential patient–clinician confrontations and to ensure a thorough exploration of the collaborative space from both the patient and clinician perspectives. All participants were provided with an introduction to the study and its aim. Informed written and oral consent was obtained from all participants before the workshops. All participants were informed that the findings are presented in an anonymous manner, compliant with ethical standards to safeguard participant confidentiality. The study was conducted in accordance with the principles outlined in the Declaration of Helsinki and by Danish law this study was considered exempt from full ethical approval.

## Results

Of 221 potential participants, a total of 28 participants were included and participated across the three workshops. Furthermore, 13 stakeholders were invited to participate in the interviews, of which six participated. In workshop 1, eight patients and two relatives participated (n = 6 females (60%), mean age 62.7±8.9, range 45-72). The patients had an average symptoms duration of 38.2±40.3 months, range 6-120). In workshop 2, ten physiotherapist and two chiropractors participated (n = 8 females (58%), mean age 36.4.7±9.3, range 26-50). On average, participants in workshop 2 had been working with this SAPS patient group for 8.9±6.5 years, range 2-24). Lastly, in workshop 3, six GPs participated (n = 3 females (50%), mean age 42.3±16.2, range 29-63). On average, participants in workshop 3 had been working with this patient group for 27.6±13.1 years, range 1- 28). From the six interviews, three were physiotherapists, one patient, one relative and one GP (n = 4 females (66%), mean age 44.6±12.7, range 33-61). Across the HCPs, they had been working with this patient group on average for 7.0±3.3 years, range 3-11).

### Results of themes within and between the workshops

Reflexive thematic analysis identified 20 initial themes and 59 subthemes across the workshops and interviews. Workshop-specific themes were subsequently compared and synthesised into two overarching categories representing (1) barriers and tensions affecting SDM in SAPS care and (2) visions for future SDM interventions. These categories comprised 11 themes that were consistently identified across stakeholder groups (**Figure 3**). Representative quotations are provided throughout this section, while the complete coding framework and supporting quotations are available in **Appendices 3**, **4** and **5**.

**Figure 3.**
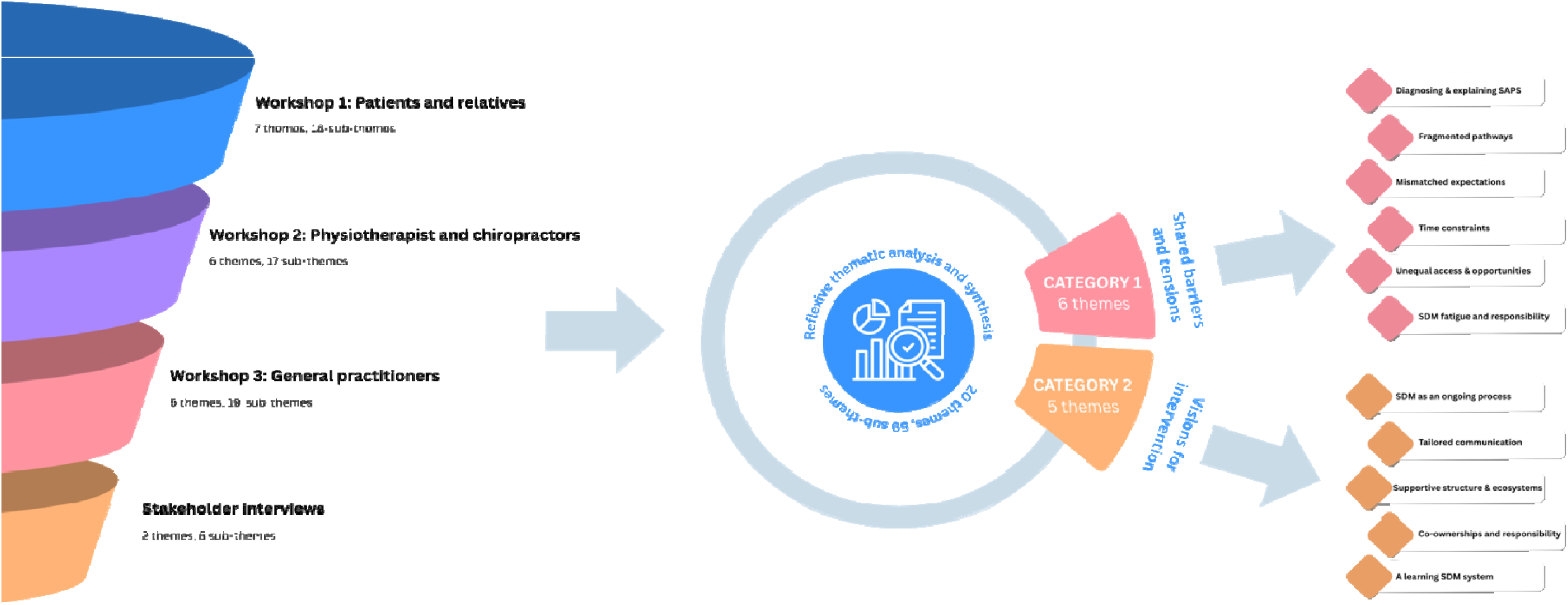
Overview of themes derived from the reflexive thematic analysis.

### Category 1: Shared barriers and tensions

Diagnostic ambiguity and inconsistent explanations of SAPS created uncertainty and reduced confidence in treatment decisions. Participants described fragmented care pathways, repeated storytelling across providers, and limited communication between professionals, resulting in poor continuity of care. Expectations regarding SDM were often misaligned, with patients seeking clearer recommendations while clinicians attempted to facilitate collaborative decision-making despite treatment uncertainty. Time constraints, inconsistent information, and both information overload and underload further limited meaningful involvement in decisions. Participants also highlighted structural inequities related to geography, finances, digital access, and health literacy, alongside concerns that SDM could become burdensome when responsibility was transferred to patients without sufficient support (See supporting quotes for category 1 themes in **Appendix 4**).

### Category 2: Visions for future SDM interventions

Participants consistently described SDM as an ongoing process rather than a single consultation. Future interventions should therefore provide tailored communication tools, support reflection before and after consultations, improve continuity across providers, strengthen digital and organisational infrastructure, facilitate shared responsibility between patients and clinicians, and establish mechanisms for continuous learning and feedback. Together, these themes emphasised that successful SDM requires coordinated support across the entire care pathway rather than isolated consultation tools (See supporting quotes for category 2 themes in **Appendix 5**).

### Matrix analysis

The matrix analysis revealed a complex interplay of tensions, divergent expectations, and systemic frictions surrounding SDM in the context of SAPS. Drawing from the three workshops and interviews, we identified how challenges operate at multiple levels and across stakeholders. The results represent isolated barriers, and demonstrated how challenges clustered across stakeholders, domains (e.g., decision-making, care pathways, and organisational structures), and healthcare system levels, providing a richer understanding of where and why SDM is hampered. At the micro level, patients described difficulties making sense of their diagnosis, the absence of a clear treatment trajectory, and the burden of being expected to make decisions without adequate information. They sought both autonomy and guidance, but found themselves overwhelmed by uncoordinated advice, time-limited consultations, and unclear responsibilities. Relatives similarly reported being left out of consultations yet still tasked with supporting decisions. These tensions were particularly acute when trust was fragile or when health literacy was low. At the meso level, the analysis identified substantial convergence regarding fragmented interprofessional communication and poor continuity across the care pathway. Physiotherapists and chiropractors described difficulties aligning evidence-based recommendations with patients’ expectations for definitive diagnoses and treatments, while GPs highlighted the constraints imposed by diagnostic uncertainty, limited consultation time, and their gatekeeping role within primary care. Divergence emerged regarding responsibility for decisions: clinicians generally viewed SDM as a collaborative process requiring active patient participation, whereas many patients expected clearer recommendations and greater professional leadership, particularly when treatment uncertainty was high. At the macro level, participants consistently identified structural barriers that constrained SDM irrespective of individual clinician behaviour. Unequal access to services across municipalities, financial considerations, fragmented digital communication systems, and organisational incentives favouring productivity over dialogue limited opportunities for consistent SDM across settings. These system-level constraints frequently amplified tensions identified at the patient and clinician levels. Integrating these findings revealed that barriers to SDM rarely occurred in isolation but instead reflected interacting tensions across system levels and stakeholder groups. This synthesis informed the conceptual model (**Figure 4**), which illustrates the relationships between actors, tasks, tensions, and contextual influences shaping SDM in SAPS care. Furthermore, the model identified actionable leverage points that directly informed the six context-specific design features presented in **Figure 5**, providing an empirically grounded foundation for developing a future SDM intervention for SAPS.

**Figure 4.**
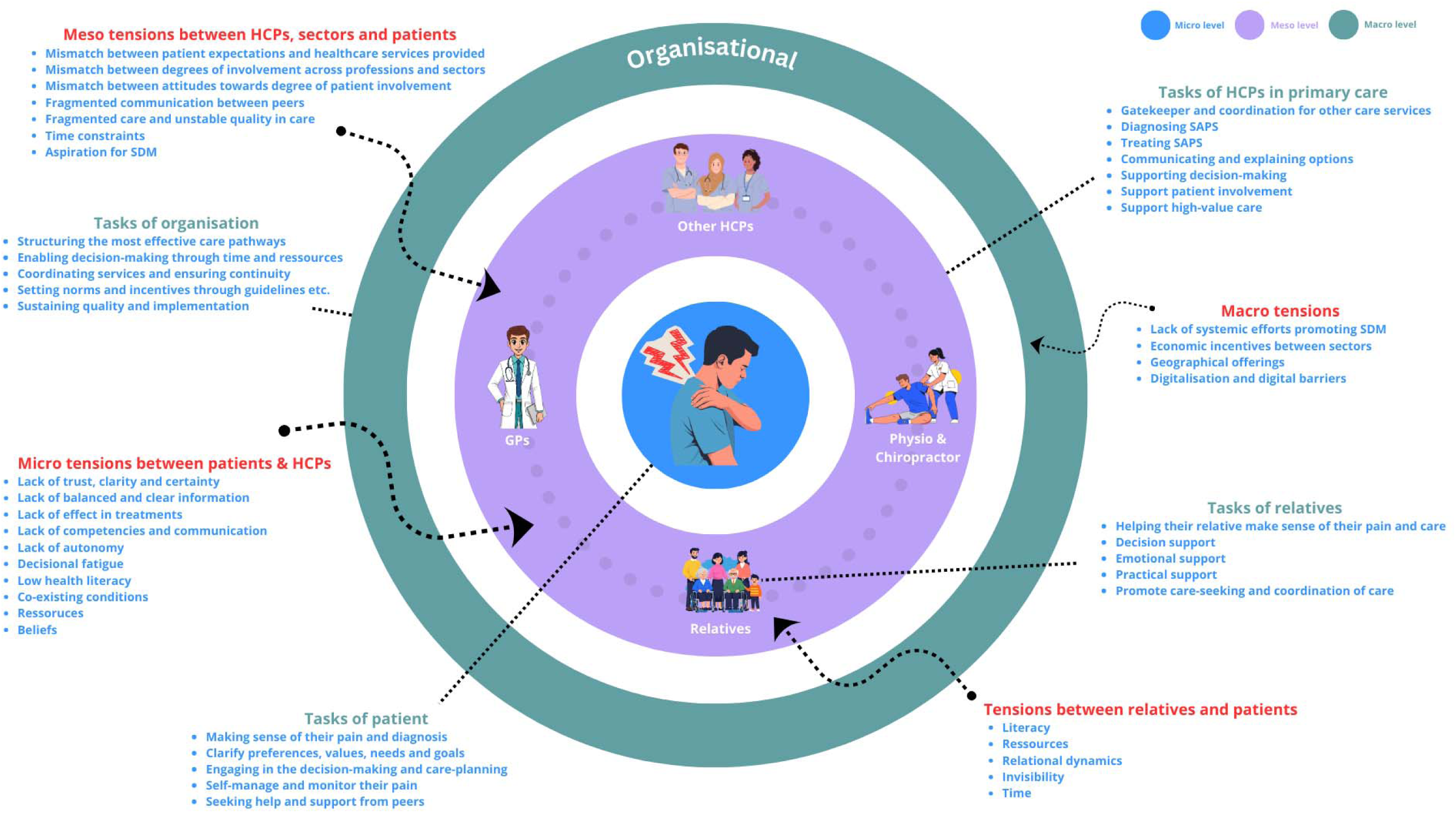
Conceptual model of SDM tensions, roles, and dynamics in the care of SAPS.

**Figure 5.**
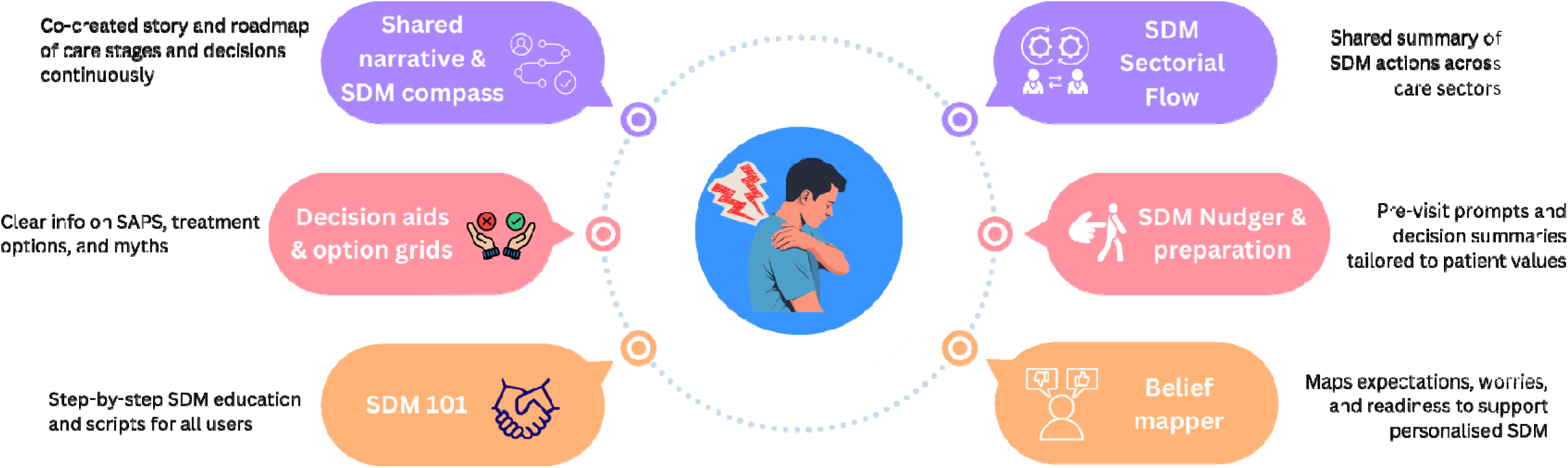
Context-specific design features for supporting SDM in the primary care management of SAPS.

### Design features for developing an SDM intervention for SAPS

Building on the matrix analysis and the visions articulated across workshops and stakeholder checks, six interlinked, context-specific design features were identified to guide development of a future SDM intervention for SAPS in primary care (See **figure 5**). First, the “*Shared narrative and SDM compass*”-feature aims to support meaning-making by co-constructing the patient’s shoulder pain story and visually outlining care stages, decision points, and goal tracking over time. Second, “*Decision Aids*”-feature addresses both information overload and underload by providing accessible explanations of SAPS, treatment options, care pathways, and tailored “myth-busting” content to manage mismatched expectations. Third, “*SDM 101*”-feature offers foundational education on what SDM is and how to practice it, including step-by-step guides, adaptive communication strategies, and mini-scripts for clinicians. Fourth, the “*SDM Sectorial Flow*”-feature is a structured summary tool that captures and communicates how SDM has been enacted during encounters, ensuring alignment across the care pathway. Fifth, the “*SDM nudger and preparation*” enables patients and relatives to reflect on values, preferences, and “what matters most” before a consultation, and includes post-consultation summaries to promote continuity. Finally, the “Belief mapper”-feature facilitates exploration of patients’ expectations, health beliefs, previous experiences, worries, and decisional readiness, offering clinicians insight into barriers that may hinder SDM.

## Discussion

Across workshops and interviews, we identified a complex web of tensions, misalignments, and structural barriers that currently constrain meaningful SDM. At the same time, participants articulated a pragmatic yet hopeful vision for a more supportive and flexible SDM process, one that is sustained over time, tailored to individual capacity, and integrated across the care pathway. These insights formed the foundation for a set of empirically grounded design features for a future SDM intervention package.

Participants described SDM as aspirational but often fragile in practice. Current SAPS care is characterised by diagnostic ambiguity, mismatched expectations, time constraints, and fragmented communication across providers (Samantha Charmaine Bengtsen et al., 2025b; Rhon et al., 2025). These findings align with prior studies showing that SDM falters when patients lack clarity about their condition, clinicians are uncertain about best next steps, and systems fail to support longitudinal dialogue or consistent messaging (Andersen et al., 2025). Importantly, many of these challenges are not specific to SAPS but reflect broader tensions in musculoskeletal and primary care, particularly when managing persistent pain when there is rarely one superior treatment (Khan et al., 2025; Storey et al., 2013). At the same time, participants pushed against the assumption that more information or more choice alone would lead to better decisions (Joseph-Williams et al., 2014). Many patients and relatives described how “*being offered a choice*” often felt like being left to carry the weight of uncertainty alone. Clinicians, too, reflected on the unintended consequences of SDM policies that promote involvement without equipping them with the time, tools, or system-level alignment to enact it meaningfully. These findings echo concerns around the lack of knowledge on the burden of SDM, including the impact of tokenism, decision fatigue, and the need for scaffolding, both cognitive and structural, to make SDM feasible in everyday care (Barone and Klatman, 2025; Montori et al., 2026). Our study extends this by showing how these issues play out across multiple care settings and by offering specific, co-developed features for a future intervention.

The future SDM envisioned by stakeholders was not framed around a singular consultation or decision point but as a distributed process unfolding over time, across roles, and within the patient’s lived context. The resulting design features, including a shared SDM compass, tailored decision aids, tools for framing and preparation, and sector-bridging communication supports, respond directly to the tensions identified in the matrix analysis. Rather than imposing a rigid protocol, the design features aim to support local adaptation, acknowledging that SDM must accommodate variation in health literacy, digital access, professional norms, and care pathways. In this sense, the intervention concept aligns with calls for adaptive, complexity-informed approaches to SDM and implementation (Butterworth et al., 2026; Clayman et al., 2024).

Several strengths of this study enhance the trustworthiness of the findings. We used an iterative, multi-method design combining structured workshops with follow-up stakeholder interviews to triangulate perspectives and refine outputs. Our use of matrix analysis allowed us to surface both converging and diverging logics across actors, and to embed these dynamics into a conceptual model that clarifies pressure points for future intervention. Furthermore, our sampling included participants from across multiple sectors, increasing the relevance of our findings to real-world practice. Nonetheless, limitations must be acknowledged. Our findings are specific to a regional Danish healthcare context and may not fully translate to other settings with different clinical structures. Moreover, while we sought to capture a wide range of voices, certain groups, such as non-Danish speakers or those with low digital literacy, have been underrepresented. Finally, although the co-developed design features are grounded in lived experience and practice-based knowledge, their feasibility and impact remain to be tested in real-world settings. Implementation and evaluation will require close attention to contextual tailoring, equity, and system integration.

This study contributes to the emerging literature on participatory intervention development in SDM by offering both a conceptual model of tensions and a concrete starting point for intervention design. By reframing SDM as a system of distributed support rather than a singular act of choice, our findings open new avenues for strengthening patient-clinician collaboration in complex care pathways. Future research should build on this foundation through pilot testing, iterative refinement, and evaluations of effectiveness and implementation that attend to context, mechanism, and outcome. In doing so, SDM interventions may be able to move from static tools alone toward dynamic, learning systems that meet the realities of patients and professionals alike.

## Conclusion

This study reframes SDM in SAPS from a discrete clinical task to a longitudinal, system-dependent process shaped by uncertainty, fragmentation, and varying patient capacities. Our findings highlight the need for integrated, context-sensitive approaches that support alignment across patients, clinicians, and care pathways. The proposed design features offer a practical foundation for developing and testing such interventions in primary care.

## Contributions

KDL is the guarantor. KDL, NEF, JLO, JLT, JS and MSR drafted the first version of the protocol. All authors contributed to the conception and design and provided critical scientific input into the protocol. KDL, responsible for the workshop and data analysis. All authors developed the data analysis & synthesis approach. All authors approved the final version of the protocol.

## Data Availability

All data produced in the present work are contained in the manuscript

## Appendix 1. Schedule of workshops

Below is an example of how the workshop swas structured.

| Time/phase | Agenda | Responsible |
| --- | --- | --- |
| **<u>Introduction</u>**<br>25 min<br>+ Breakfast | Welcome and introduction to the research project (project aim, goal and agenda) and participants.<br>Presentation (inspiration)<br>Summary and questions. | **KDL** |
| 5 min | **Break** |  |
| **<u>Critique:</u>**<br>45 min | Introduction and instructions on exercises and how to complete design card exercises. Description of the case: Plenum. (5 min)<br>Card exercises: (Domain cards and Problem cards)<br>Red post-its. (30 min)<br>**Approach:**<br>- Participants read the case aloud.<br>- By using the domain cards, participants create a narrative outlining the barriers related to shared decision-making in clinical practice (Blank cards fill gaps).<br>- Participants select problem cards and place them adjacent to the barriers, illustrating the problems with managing the patients in the current healthcare system<br>- Participants use the red post-its to describe ‘why’ they see this as a problem. (insights)<br>Insights are selected and presented to other groups (ranking) (10 min) | **KDL** |
| 10 min | **Break** |  |
| **<u>Vision:</u>**<br>45 min<br>(visions) | Introduction and instructions on exercises and how to complete design card exercises. (5 min)<br>**<u>Framing “all is possible, except for what is impossible”</u>**<br>Introduction of solutions cards and initial thought about the design of the combined care pathway. (30 min.)<br>**Approach:**<br>- Participants familiarize themselves with the different solutions on the solutions card.<br>- Participants select solutions cards and place them next to the relevant problem cards.<br>- Using the yellow post-its, they write how this solution could prove valuable to them.<br>Discussions (plenum)<br>- Each group selects two solutions and present them to get feedback (ranking) (10 min) | **KDL** |
| 10 min | **Break** |  |
| **<u>Reality:</u>**<br>30 min. | The implementation phase is introduced, along with the exercises and an introduction to storyboarding. (5 min.)<br>-Participants are asked to choose two specific solutions, which they are asked to explore in depth. | **KDL** |
|  | - Values are noted on green post its.<br>- Paper, crayons and pictures are provided and the participants are asked to sketch storyboards. (10 min.)<br>-The session will be concluded with a presentation of the visions and the storyboards of each group in plenum and feedback including ranking.<br>-The facilitator notes visions and values on whiteboards as we go along. (15 min.) |  |
| **<u>Summary</u>**<br>5 min. | Questions, practicalities and handing in permission to record data. (5 min.) | **KDL** |

## Appendix 2. Case vignette

Our fictious person is a 52-year-old office worker who lives in Aalborg with her partner. She visits her general practitioner after experiencing increased shoulder pain over the past five months. She doesn’t recall any specific injury, but says the pain has gradually worsened and is now affecting her sleep, daily activities, and ability to keep up with household chores and her part-time job.

Her general practitioner performs a clinical examination and is told that she has probably overloaded it. She is referred to physiotherapist and receives a pamphlet about exercises. She is also offered the option of imaging but is told it may not change the treatment plan.

At the physiotherapy clinic, she expresses frustration about not having a clear diagnosis and is unsure whether the exercises are helping. After six weeks of supervised sessions (with a physiotherapist), her symptoms improve slightly but remain bothersome. She asks both her physiotherapist and general practitioner whether she should consider a cortisone injection or be referred to an orthopaedic specialist. The professionals offer different views, one recommends continuing physiotherapy, another thinks an injection may be reasonable or waiting a bit longer.

She feels confused. She wants to avoid unnecessary treatment but also feels the pain is affecting her well-being. She is unsure how to make the right decision and would like more clarity and support in choosing the next step.

## Appendix 3: Overview of Themes and Sub-Themes by individual workshops

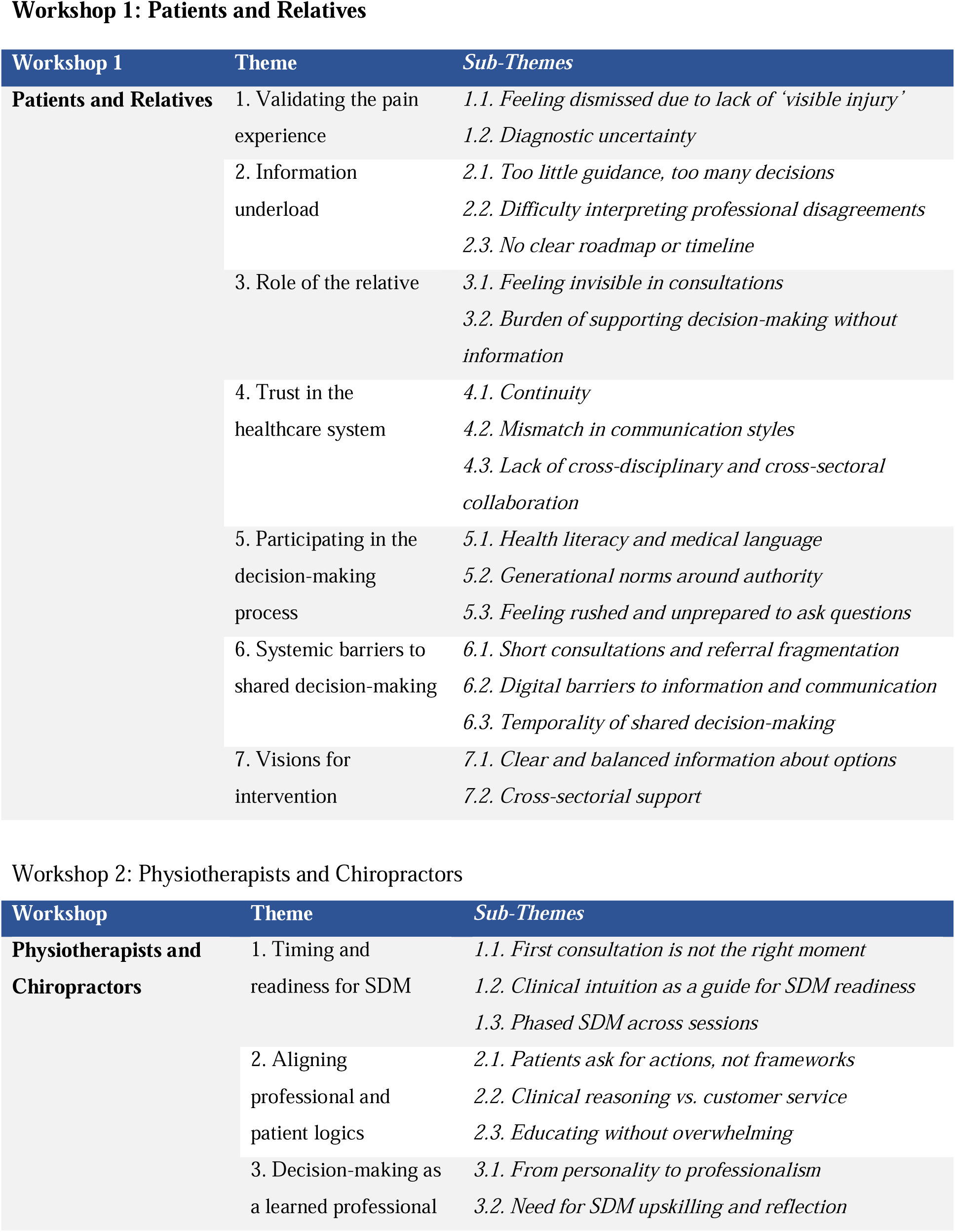
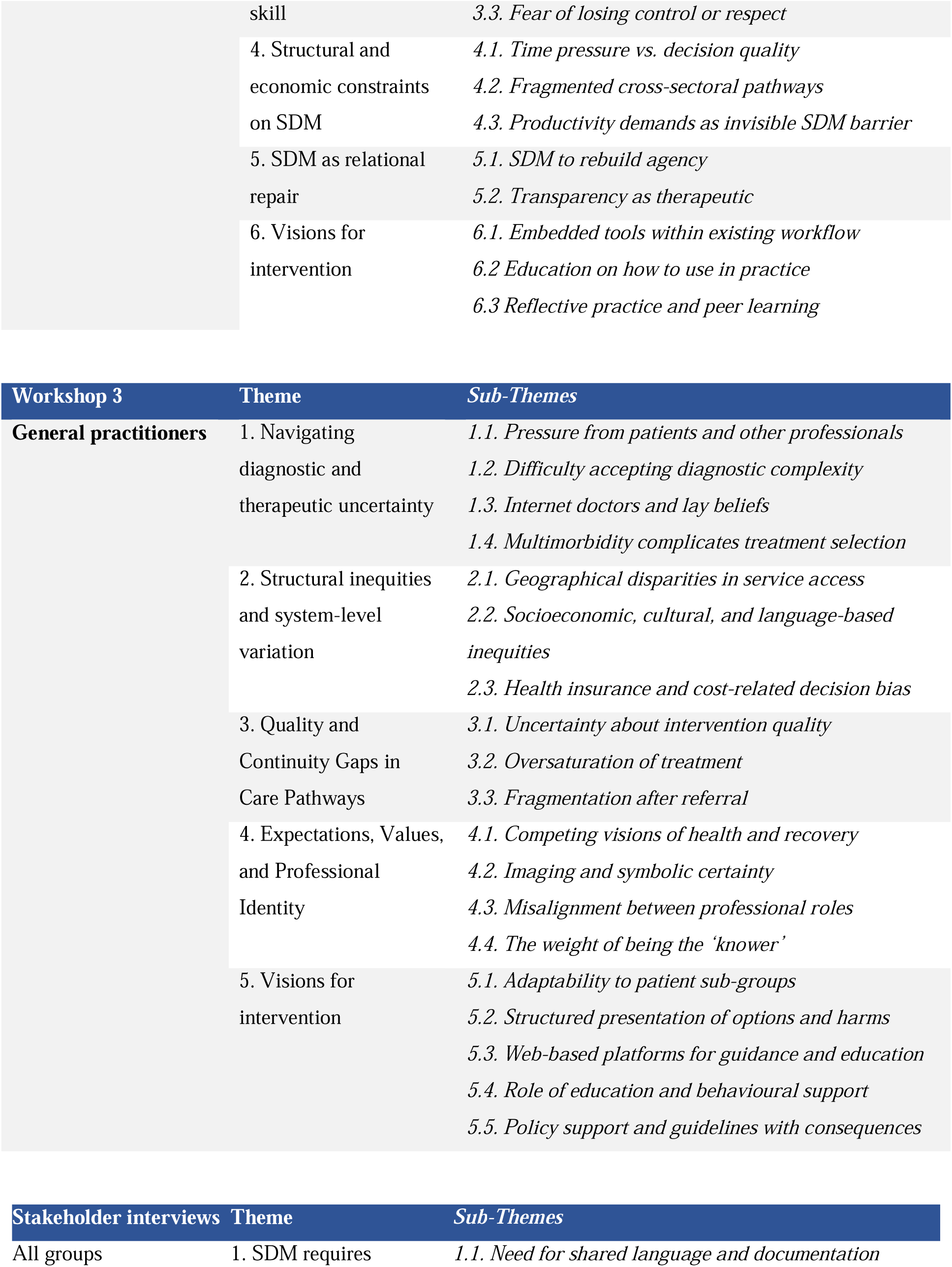
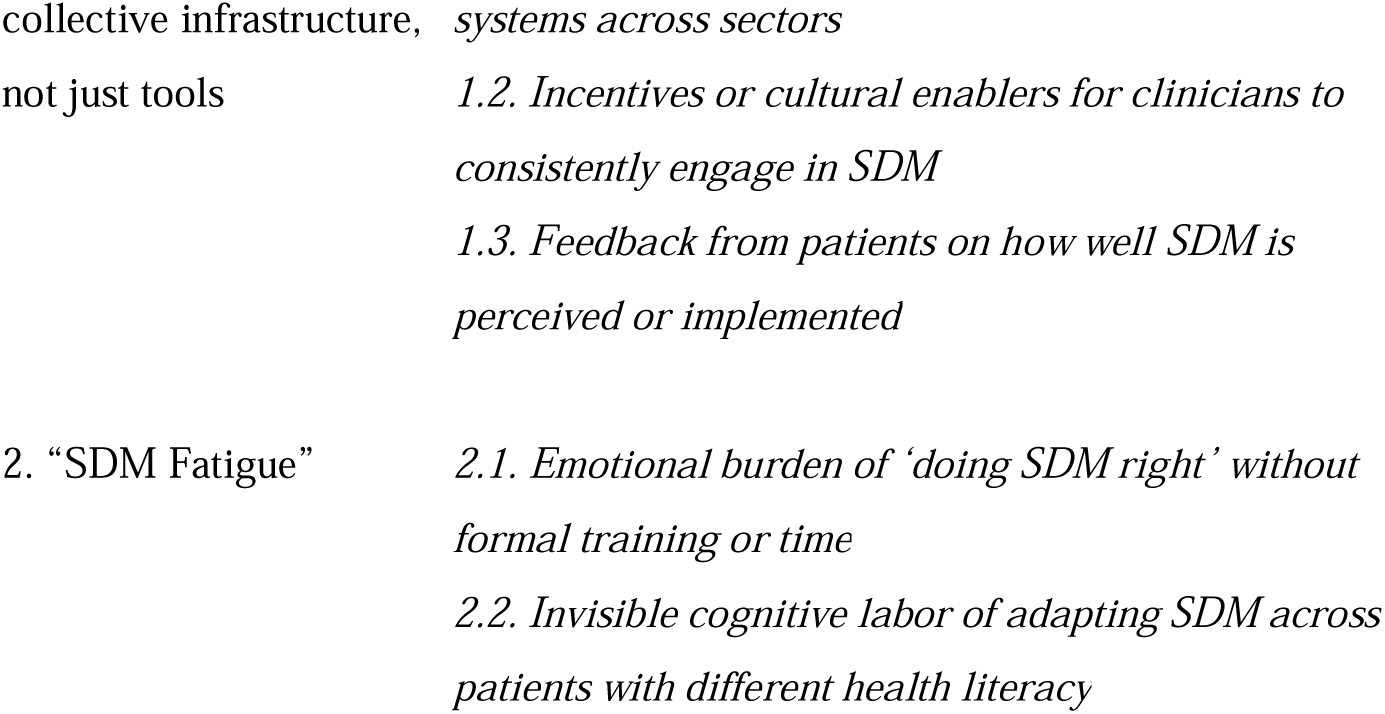

## Appendix 4: Supporting quotes for category 1

### Category 1: Shared barriers and tensions in SDM for SAPS

#### Theme 1: Diagnosing, Naming and Explaining SAPS

This theme highlights how uncertainty surrounding the diagnosis of SAPS challenges the current approaches to clinical management and understanding of SAPS, and participation in SDM. While some HCPs saw SAPS (or similar label such as rotator cuff related shoulder pain) as a useful label for a clinical presentation, many patients and relatives experienced the label (and similar labels) as vague and sometimes unhelpful. Patients described the label as something that was difficult to understand and explain to others. They associated the diagnostic uncertainty with emotional unease, often feeling they had to advocate for further tests, particularly imaging, to legitimise their experience. As one patient shared,

> *“When you’re told its SAPS, but apparently no one really says what that means, you start to wonder if it’s even real. Also, it makes me question it even more, when I hear different labels from different doctors and physios” [Workshop 1 – Patient]*

HCPs acknowledged that the SAPS diagnosis offers limited guidance. For GPs the label was sometimes used as a placeholder in referral letters, rather than a precise diagnostic statement. This was further reflected on by a physiotherapist:

> *“It’s a category we fall back on, but it doesn’t help me much saying and knowing what will help the person in front of you.” [Workshop 2 – Physiotherapist]*

This diagnostic uncertainty was further exacerbated by inconsistent messaging across the care pathway. Participants described receiving different explanations from GPs, physiotherapists, and chiropractors, each with varying degrees of certainty or contradiction. These inconsistencies eroded trust and made it harder for patients to engage in SDM, particularly when expected to weigh options without a shared understanding of what the condition entailed. Stakeholders described how SAPS is often explained in technical or biomechanical terms (e.g., impingement, bursitis, tendonitis), but these labels could create more confusion than clarity. A GP acknowledged:

> *“We use words that sound like diagnoses but are just guesses and poor explanations. I find it hard to explain this issue, when all that the patient wants is certainty and precise guidance.” [Workshop 3 – GP]*

Several participants across groups linked the vagueness of SAPS to a broader dilemma in musculoskeletal care: the tension between clinical uncertainty and the need to act. While few HCPs embraced this uncertainty as an opportunity for collaborative exploration, others felt pressure to simplify the message or offer interventions prematurely.

#### Theme 2: Fragmented Pathways and Loss of Continuity

Participants across all stakeholder groups recognized that the SAPS care pathway often could be perceived as uncoordinated and difficult to navigate. Patients and relatives reported feeling “passed around” between providers and sectors without clear communication, follow-up, or a sense of overarching responsibility. This fragmentation weakened trust, delayed decision-making, and left patients and relatives unsure of what came next and their expectations. Patients commonly described a feeling of “starting over” each time they transitioned between care providers. One patient shared:

> *“I had to tell my story again and again. Every time, from scratch. Often, I never really knew if anyone knew what had happened before. I even had to explain it to the same person several times – the least I can expect is that they will find time to look up key information in my journal” [Workshop 1 – Patient]*

Relatives also expressed concern over this lack of continuity, especially when trying to support decision-making on behalf of the patient. A relative reflected:

> *“We were told to just wait or try something—but there was no one who seemed to follow the whole process.” [Workshop 1 – Relative]*

HCPs also expressed frustration with the health care system’s fragmentation. GPs noted the absence of feedback loops after referring patients to physiotherapists or specialists (such as orthopaedic surgeons). One GP commented:

> *“I send someone for physio, but then I don’t hear anything. I have no idea if they improved, dropped out, or got referred further. Also, I have no idea how the quality of the treatments was. Sure – exercise can be good, but I don’t know exactly how it was done, what doses and so on” [Workshop 3 – GP]*

Physiotherapists and chiropractors described uncertainty about what advice patients had already received or what expectations were set in previous encounters, mostly from the patients’ GP. This not only complicated treatment planning but also undermined attempts to engage patients in SDM at later stages. A physiotherapist explained:

> *“Sometimes we contradict each other without knowing it. That’s not fair to the patient. I also experience that it can be difficult to combat other HCPs who might not be updated on the newest knowledge” [Workshop 2 – Physiotherapist]*

Stakeholders pointed to several structural causes, including the separation of primary and municipal services, variation in available local offers (e.g., rehabilitation or imaging access), and inconsistent digital communication tools. Digital handoffs between systems were seen as partial solutions but not yet capable of supporting a coherent care experience. Decisions made in one setting often become disconnected from the next, leaving patients with the burden of coordination. As one patient from the interviews noted,

> *“I often think that a doctor should not only be good at making decisions, but they also need to make sure someone carries them forward.” [Workshop 1 – Patient]*

Interestingly, this was also contradicted by other patients, who saw the lack of control by HCPs as an invitation to involve themselves in their own care:

> *“I actually think it is okay that my doctor is not always the one leading. I have resources and I want to have the feeling that I’m in charge of what I want. In such situations where I feel “invited”, I also engage much more in the conversation and ask questions.” [Workshop 1 – Patient]*

Improving continuity of care, clarifying roles across sectors, and strengthening communication infrastructure were seen as essential foundations for any SDM intervention aiming to support patients through the entire SAPS care trajectory.

#### Theme 3: Mismatched expectations and communication styles

This theme captures how differing expectations and communication habits between patients, relatives, and clinicians can complicate the SDM process for SAPS. Across workshops, participants described misalignment in goals, language, and assumptions, which could hamper decision-making. Most patients and relatives approached consultations expecting clear recommendations and expert guidance. When instead presented with choices, especially with uncertainty of their effectiveness, they felt unprepared or overwhelmed. One patient reflected:

> *“I didn’t want to be in charge, I wanted to understand and be understood. But the way the doctor explained it, it felt like I had to pick something when I didn’t even know what the real problem was. To be honest, this turned out worse when I went on in the system and I was told something entirely different” [Workshop 1 – Patient]*

For relatives, this ambiguity created a secondary burden of interpreting medical language and supporting decisions without feeling fully informed. Several relatives describe this communication gap as trying to solve a puzzle with missing pieces. HCPs were often aware of these tensions in communication but struggled to resolve them in practice. GPs described trying to balance professional caution with patient expectations for clarity or action. A chiropractor noted:

> *“People come wanting answers, and we can’t always match that – we most often also come with some sort of uncertainty in the management, but also in knowing what this patient has been told before. It’s often an odd situation where you are not sure if explaining yourself and what others have thought is meaningful” [Workshop 2 – chiropractor]*

Physiotherapists echoed this concern, describing how some patients perceived neutral or evidence-informed recommendations as disinterest or indecisiveness. One said:

> *“When I say, ‘we can try this or that,’ I mean it genuinely. But some hear it as ‘I don’t know’ or ‘it doesn’t matter.’” [Workshop 2 – Physiotherapist]*

Cultural norms also played a role. Some patients were hesitant to question the HCP or initiate dialogue, feeling that this would be seen as disrespectful or confrontational. In contrast, HCPs expected active engagement as a prerequisite for SDM. A physiotherapist observed:

> *“Some patients expect us to lead completely, while I want to involve them. That can create this awkward gap, we I often think I’m looking a bit incompetent – I wish SDM was more accepted in the general public” [Workshop 2 – Physiotherapist]*

This mismatch was especially visible when discussing imaging results, diagnoses, or when expectations had been shaped by previous clinicians or online information. Stakeholders noted that differing communication styles and assumptions about roles can reinforce misunderstandings, leave uncertainty unaddressed, and weaken decision quality.

#### Theme 4: Time constraints and information over- and underload

Time was consistently identified as barriers to SDM by participants across all workshops. Patients, relatives, and clinicians described how the limited duration of consultations, particularly in general practice, left insufficient room to explore options, ask questions, and engage in meaningful dialogue. At the same time, the volume and complexity of information patients were expected to absorb and act upon contributed to feelings of confusion and pressure. Patients described consultations as rushed and overwhelming, particularly when multiple decisions had to be made without time to process their meaning or implications. One patient reflected:

> *“You go in expecting a conversation, but before you even understand what’s going on, you’re being asked what you want to do. Me and my husband calls it speed dating with your own health.” [Workshop 1 – Patient]*

Relatives echoed this concern, noting how the pace of interaction often prevented them from stepping in or supporting the patient effectively. They described struggling to remember what had been said or knowing when it was appropriate to ask questions. One relative explained:

> *“It goes so fast, and if you’re not used to the language or the system, you don’t even know what to ask. You just nod and leave more confused than when you came in.” [Workshop 1 – Relative]*

HCPs, too, acknowledged the impact of limited time on their ability to practise SDM. GPs described being caught between the need to cover medical basics, dealing with the complexity of SAPS, and expectations for involvement. A GP remarked:

> *“We want to include people, but 15 minutes is 15 minutes. By the time you’ve heard their story and done a basic exam, you’re almost out of time. I wish I could do more to facilitate SDM, but I think it is difficult with the time I have” [Workshop 3 – GP]*

Physiotherapists described similar tensions, especially when patients arrived with expectations already shaped by previous encounters or online information. They noted that time was not just needed for explanation but also for recalibration. One said:

> *“Often I spend the first session just trying to make sense of what they’ve already been told. It’s not about deciding yet, it’s about getting on the same page.” [Workshop 2 – Physiotherapist]*

Information over- and underload, was a recurring theme, mostly highlighted by patients and relatives. Participants across groups described situations in which patients were asked to weigh options or decide on care pathways despite feeling ill-equipped to do so. Across all workshops, participants emphasised the value of interventions that support SDM before and after consultations. Similarly, several stakeholders suggested that decision-making aids and visual tools could help the patients structuring all the information. Contrary to the patients who felt an information overload, there was also multiple patients, who experienced information underload. As one patient describes it:

> *“The longer I have had pain, the less I feel that I know and the same goes for my doctors. Right now, I’m in a state where I get no information, there’s no initiatives for me, and I kind of just feel that I drift across sectors and professionals and nobody knows anything” [Workshop 1 – Patient]*

#### Theme 5: Uneven Access, Unequal Opportunities

This theme highlights how structural inequities, across geography, socioeconomic status, educational background, and health insurance coverage, shape who can meaningfully participate in SDM and who cannot. Participants across all workshops described how unequal access to care options, varying health literacy, and financial barriers could limit the scope and quality of decisions, regardless of intention. Patients reported being offered different treatments depending on where they lived or which professional they encountered first. Several noted that “choice” was only theoretical when geography, cost, or lack of local services restricted what was actually feasible. One patient reflected:

> *“They say you can choose, but if one option is private and costs, let’s say, 500 kroner, and the other is free but has a waiting list, it’s not a real choice.” [Workshop 1 – Patient]*

Relatives echoed this, where some services (e.g., certain rehabilitation offers or imaging) were unavailable. They pointed to difficulties navigating different offers across municipalities and the private sector which led to confusion about what was covered by insurance and what was not. HCPs confirmed that such disparities shaped both what they recommended and how they framed choices. A physiotherapist commented:

> *“I have worked across different regions, and in some municipalities, I have had a good collaboration with rehab teams. In others, it’s nothing. So even if we want to co-decide, we’re not offering the same things everywhere. I think this is a big problem and I honestly I feel sorry for my old patients where the offers don’t match elsewhere” [Workshop 2 – Physiotherapist]*

GPs described the challenge of balancing realistic options with what was ideal, especially for patients with limited economic or digital resources. One GP explained:

> *“You can’t offer everyone the same plan if half of them can’t access it or understand it. Then it becomes unethical to ‘involve’ them in a decision they can’t follow through on.” [Workshop 3 – GP]*

Low health literacy was seen as another key barrier to equitable SDM. Participants discussed how patients with fewer educational resources or limited familiarity with medical systems were less likely to ask questions, seek clarification, or express preferences. Several clinicians described needing to “guess” the patient’s level of understanding or adapt their language without knowing if it helped. Stakeholders suggested that future SDM interventions for SAPS must explicitly address such inequities. Ideas included universal access to visual tools, reimbursement models that supported equal offerings, and design strategies sensitive to literacy, language, and digital divides.

#### Theme 6: SDM fatigue and the burden of responsibility

While SDM was generally supported in principle, patients, relatives, and clinicians raised concerns about the unintended emotional and cognitive burdens it can place on individuals, particularly in the context of clinical uncertainty. This theme captures how SDM, when poorly implemented or unsupported, may overwhelm rather than empower, leading to fatigue, frustration, and disengagement. Patients described feeling ill-equipped to take on decision-making responsibilities when they lacked sufficient knowledge, confidence, or certainty. Several recounted being asked to “choose” between options they didn’t fully understand, especially when clinicians appeared uncertain themselves. One patient shared:

> *“I was told, ‘you can do exercise, injection, or wait.’ But I didn’t know what any of that really meant. It felt like a test I didn’t study for.” [Workshop 1 – Patient*]

Some patients appreciated being included in the process, but only when it was accompanied by guidance and reassurance. Others felt abandoned or confused in the process, as one relative expressed concern that SDM was being used to shift responsibility away from clinicians:

> *“It’s fine to have a say, but sometimes it felt like they didn’t want to be responsible if it didn’t work. Especially, when we are having a hard time with multiple things. I think in that situation it felt a bit scary to feel a type of responsibility, when all we wanted was someone who could help and take charge of my wife” [Workshop 1 – Relative]*

HCPs also voiced this ambivalence. Several physiotherapists, chiropractors, and GPs described how SDM could inadvertently overload patients, especially those with low health literacy, multiple conditions, or psychological distress. A physiotherapist explained:

> *“Some people want involvement, but others just want clarity. You have to know when SDM helps and when it just adds weight.” [Workshop 2 – Physiotherapist]*

GPs described the challenge of navigating between empowerment and burden. One remarked:

> *“It’s not always helpful to give people a menu when they’re drowning. Sometimes they need someone to throw them a rope first – unfortunately we don’t always “get” that” [Workshop 3 – GP]*

Stakeholders noted that without time, tools, or scaffolding, SDM can become performative rather than meaningful. Patients may be invited to choose without having the support needed to weigh trade-offs or consider implications. Clinicians may feel caught between policy expectations and clinical realities. This theme suggests that SDM fatigue arises when the process is decoupled from support, when decisions are asked for but not prepared for.

## Appendix 5: Supporting quotes for category 2

### Category 2: Visions and features for Future SDM Interventions

#### Theme 7: SDM as an ongoing process, not a one-off task

This theme captures a shared vision across stakeholder groups that SDM in SAPS should be viewed not as a single event or conversation, but as a continuous, evolving process. Participants expressed a need for flexibility, revisiting decisions over time, and adapting care plans as patients’ experiences, goals, or symptoms change. Patients described how their understanding of SAPS and their treatment preferences developed gradually, often shifting after trying a particular intervention, talking to peers, or simply living with the condition over time. One patient shared:

> *“At first, I just wanted the pain gone. But after the injection didn’t work, I realised I needed to understand it better. I had new questions. But no one followed up.” [Workshop 1 – Patient]*

Relatives echoed this, noting how their ability to support the patient changed as new decisions emerged or previous ones proved ineffective. They emphasised the value of check-ins and opportunities to re-engage with professionals along the care journey. Clinicians also supported this dynamic view of SDM. Several physiotherapists and GPs highlighted how decisions often unfold across multiple sessions or shift entirely after the patient has lived with the implications of an earlier choice. A GP noted:

> *“People come back with different thoughts than when we first spoke. That’s not failure, that’s normal. We need to leave room for that.” [Workshop 3 – GP]*

Physiotherapists highlighted that patients may need time to reflect before actively participating. One explained:

> *“Sometimes the real SDM happens in the second or third session, once the trust is there, and they’ve had time to think about what matters to them.” [Workshop 2 – Physiotherapist]*

Both HCPs and stakeholders saw value in formalising SDM as a process, through checklists, follow-up calls, or digital tools that prompt ongoing reflection. Several emphasised that expectations should be set early: SDM doesn’t mean locking in a permanent choice but working together over time.

#### Theme 8: Tailored communication and framing tools

Participants across all groups emphasised the need for communication strategies and tools that adapt to patients’ individual capacities, preferences, and circumstances. Rather than relying on standardised verbal explanations, many envisioned an SDM process enriched by tailored visuals, analogies, structured overviews, and accessible digital supports. Patients frequently described difficulties grasping abstract or unfamiliar medical terms, especially when explanations were rushed or inconsistent. Several expressed a desire for visual aids to help “see” what was going on. One patient explained:

> *“I need to see it to understand it. When they talk about tendons or inflammation, I’m lost, but if I see an image or a drawing, it clicks.” [Workshop 1 – Patient]*

Relatives similarly reported struggling to retain or interpret what was said during consultations. Some suggested simple one-page handouts or traffic-light systems to summarise pros and cons of treatment options. Others wanted something to take home and discuss together:

> *“Even just a picture or a list to take home would help. It’s hard to support someone when you’re also trying to make sense of it all.” [Workshop 1 – Relative]*

HCPs described a lack of ready-to-use tools that supported SDM in the context of SAPS. Many physiotherapists and chiropractors had developed their own visuals or metaphors to explain options but noted variation across practitioners. A physiotherapist shared:

> *“You end up making your own materials. But it’s time-consuming, and not everyone is comfortable doing that. It also hard to stay updated and keep this visual updated” [Workshop 2 – Physiotherapist]*

Several GPs highlighted the need for framing strategies to help patients make sense of uncertainty without feeling abandoned. One GP explained:

> *“We need to normalise uncertainty in a way that still feels reassuring. It’s not about saying ‘I don’t know’, it’s about saying ‘here’s what we know, and here’s how we can move forward together.’” [Workshop 3 – GP]*

Stakeholders envisioned a library of optional tools, simple animations, diagrams, analogies, or digital decision aids, that clinicians could flexibly use or adapt depending on patient needs. The emphasis was not on standardisation, but customisation: tools that help structure the conversation while preserving space for personalisation.

#### Theme 9: Supportive structures and digital ecosystems

This theme reflects participants’ shared recognition that successful SDM in SAPS care is not solely dependent on individual communication but requires supportive structural and technological systems. Across workshops and stakeholder interviews, participants identified opportunities for embedding SDM into everyday workflows, digital infrastructures, and interprofessional routines, while cautioning against overcomplication or adding administrative burden. Patients and relatives emphasised that timing and delivery of information were crucial. Many wanted access to relevant materials before a consultation, to reflect and prepare. One patient explained:

> *“If I could read or watch something at home before the appointment, I’d be way more ready. Otherwise, it’s too much at once.” [Workshop 1 – Patient]*

Several suggested a central website or app that offers credible, easy-to-understand information, a catalogue of exercises, and FAQs which importantly should be something trustworthy they could return to. Relatives valued being included in this access, helping them support their loved ones. HCPs likewise expressed a strong desire for infrastructure that supports, rather than duplicates, their existing practice. GPs described wanting brief, integrated overviews within the electronic medical record, including prompts or checklists for SDM conversations. A GP shared:

> *“If we’re going to do this properly, it has to be built in, not something extra we print or remember. It should pop up when it’s relevant.” [Workshop 3 – GP]*

Physiotherapists and chiropractors envisioned digital tools that could structure initial assessments and track patient-reported outcomes, allowing shared tracking and decision-making over time. A physiotherapist noted:

> *“If we could see trends together, like pain improving, function going up, that could guide what we choose next, together.” [Workshop 2 – Physiotherapist]*

Stakeholders also highlighted the need for cross-sectoral alignment. Disconnected digital systems between general practice, municipal rehab, and private physiotherapy were seen as barriers to continuity and collaborative care. Several suggested that future SDM interventions should include shared documentation platforms or at minimum, a way for patients to carry their decisions and information across providers. Importantly, participants cautioned against digital overload. Tools should be simple, optional, and adaptable. Digital equity and literacy was a concern: some patients lack access, skills, or trust in digital health.

#### Theme 10: Co-ownership and rebalancing responsibility

Participants envisioned a future SDM model for SAPS that promotes a more balanced distribution of responsibility between patients, clinicians, and the wider system. Rather than shifting the decision-making burden onto any one party, they advocated for co-ownership, where roles are clearly defined, support is equitably distributed, and the process of arriving at a decision is shared, not outsourced. Patients and relatives repeatedly described the tension between being offered a choice and feeling solely responsible for the outcome. Some interpreted SDM as a form of delegation rather than collaboration. One patient explained:

> *“It’s not that I don’t want a say, but it felt like I had to make the decision alone, and then live with it if it went wrong.” [Workshop 1 – Patient]*

Relatives echoed this concern, particularly when they were asked to “support” the patient without having access to the necessary information or understanding. They wanted their roles acknowledged, not as decision-makers per se, but as partners who could help the patient reflect and follow through. Clinicians, meanwhile, described the difficulty of navigating responsibility in a system that increasingly emphasises patient autonomy. A GP noted:

> *“There’s this grey zone now, are we guiding, recommending, or just listing options? Patients want help, but we’re also told not to steer too much.” [Workshop 3 – GP]*

Physiotherapists and chiropractors raised similar concerns. While many had embraced a more patient-centred approach, they felt there was still a lack of structure around how responsibility is negotiated. One physiotherapist reflected:

> *“Sometimes I sense the patient wants me to decide, and other times they want full control, but we rarely talk about it directly.” [Workshop 2 – Physiotherapist]*

Stakeholders highlighted the need for an SDM model that actively facilitates role clarification. This includes tools to help patients reflect on how much involvement they want, prompts to guide clinicians in exploring this preference, and support systems to ensure decisions are followed through collaboratively. Some participants also emphasised that co-ownership extends beyond the consultation. For SDM to work, the healthcare system must share responsibility by ensuring equitable access to follow-up, providing clear guidance, and avoiding contradictory messages across providers.

#### Theme 11: Building a learning SDM system

Across all workshops and stakeholder interviews, participants articulated a future vision where SDM in SAPS is not treated as a static protocol, but as a learning system, constantly improving based on feedback, real-world use, and evolving clinical and patient needs. This theme emphasises the importance of reflection, evaluation, and adaptive infrastructure in ensuring SDM becomes a sustainable and responsive part of SAPS, and musculoskeletal care in general. Patients and relatives described frustration with one-size-fits-all recommendations and saw value in tools or systems that could learn from their feedback. Several expressed the desire for a more iterative care process, one where earlier decision could be revisited and adjusted as outcomes unfolded. One patient remarked:

> *“We tried something, but when it didn’t work, there was no next step. I wish there was a system that asked: ‘how did it go?’ and helped us figure out what to do next.” [Workshop 1 – Patient]*

Relatives wanted more structured follow-up mechanisms, not just to check on treatment progress, but to reflect on whether the decisions made still aligned with the patient’s goals and values. HCPs echoed this process and noted that SAPS management often involves trial and error, yet the system rarely supports revisiting or learning from those trials. A physiotherapist explained:

> *“We don’t really capture what happens after the choice is made. Did it work? Why or why not? That data could help all of us improve.” [Workshop 2 – Physiotherapist]*

GPs described how a lack of shared learning across sectors or settings limited their ability to refine their approach to SDM. One GP noted:

> *“We make decisions in isolation, and then the patient disappears into another part of the system. There’s no going back, no way to learn if what we did helped.” [Workshop 3 – GP]*

Stakeholders proposed that a “learning SDM system” should include mechanisms for patient feedback, peer learning among HCPs, and integration of decision outcomes into quality improvement initiatives. Doctors suggested dashboards or registries that allow for population-level monitoring of SDM processes, what was offered, chosen, and with what result. Importantly, participants cautioned that such a system must respect privacy, be easy to use, and avoid becoming another administrative burden. Its purpose should be to “close the loop” and not just to track choices, but to inform better ones in the future.

## References

Andersen, L.N., Waddell, A., Boland, L., Rathleff, M.S., Hansen, M.P., Thomsen, J.N.L., Elwyn, G., Jepsen, J.F., Lyng, K.D., 2025. Implementation of Shared Decision-Making in the Management of Chronic Musculoskeletal Pain: a scoping review. medRxiv 2025.10.02.25336876. 10.1101/2025.10.02.25336876

Barone, M.T.U., Klatman, E., 2025. A Seat at the Table, But on Whose Terms? The Illusion of Meaningful Engagement. J. Patient Exp. 12, 23743735251395370. 10.1177/23743735251395370

Bengtsen, Samantha C., Rathleff, M.S., Zadro, J.R., Olesen, J.L., Foster, N.E., Thomsen, J.L., Elwyn, G., Søndergaard, J., Lyng, K.D., 2025. Impact of a Decision Aid on Perceptions of Shared DecisionlMaking in the Primary Care Management of Patients With Subacromial Pain Syndrome: A TwolPhased MultilMethods Study. Musculoskelet. Care 23, e70172. 10.1002/msc.70172

Bengtsen, Samantha Charmaine, Zadro, J.R., Rathleff, M.S., Foster, N.E., Thomsen, J.L., Olesen, J.L., Søndergaard, J., Lyng, K.D., 2025a. Exploring the decisional needs of patients living with subacromial pain syndrome: A qualitative needs assessment study. Musculoskelet. Sci. Pract. 76, 103255. 10.1016/j.msksp.2025.103255

Bengtsen, Samantha Charmaine, Zadro, J.R., Rathleff, M.S., Foster, N.E., Thomsen, J.L., Olesen, J.L., Søndergaard, J., Lyng, K.D., 2025b. Exploring the Decisional Needs of Patients living with Subacromial Pain Syndrome: a qualitative needs assessment study. Musculoskelet. Sci. Pr. 103255. 10.1016/j.msksp.2025.103255

Birt, L., Scott, S., Cavers, D., Campbell, C., Walter, F., 2016. Member Checking. Qual. Heal. Res. 26, 1802–1811. 10.1177/1049732316654870

Braun, V., Clarke, V., 2006. Using thematic analysis in psychology. Qualitative Research in Psychology 77–101.

Bruch, J.D., Khazen, M., Mahmic-Kaknjo, M., Légaré, F., Ellen, M.E., 2024. The effects of shared decision making on health outcomes, health care quality, cost, and consultation time: An umbrella review. Patient Educ. Couns. 129, 108408. 10.1016/j.pec.2024.108408

Butterworth, J.E., Mattick, K., Richards, S.H., 2026. It is time to recognise shared decision-making as a complex intervention. Patient Educ. Couns. 144, 109447. 10.1016/j.pec.2025.109447

Clarke, V., Braun, V., Frith, H., Moller, N., 2019. Editorial Introduction to the Special Issue: Using Story Completion Methods in Qualitative Research. Qual Res Psychol 16, 1–20. 10.1080/14780887.2018.1536378

Clayman, M.L., Elwy, A.R., Vassy, J.L., 2024. Reframing SDM Using Implementation Science: SDM Is the Intervention. Méd. Decis. Mak. 44, 859–861. 10.1177/0272989x241285418

Diercks, R., Bron, C., Dorrestijn, O., Meskers, C., Naber, R., Ruiter, T. de, Willems, J., Winters, J., Woude, H.J. van der, Association, D.O., 2014. Guideline for diagnosis and treatment of subacromial pain syndrome. Acta Orthop 85, 314–322. 10.3109/17453674.2014.920991

Elwyn, G., Frosch, D., Rollnick, S., 2009. Dual equipoise shared decision making: definitions for decision and behaviour support interventions. Implement Sci 4, 75. 10.1186/1748-5908-4-75

Elwyn, G., Gulbrandsen, P., Leavitt, H., Abukmail, E., Clayman, M.L., Edwards, A., Finderup, J., Fisher, A., Grande, S.W., Hahlweg, P., Hoffmann, T., Hou, W.-H., Hernández-Leal, M.J., Leung, D., Lu, W., Mandelkow, L., Pecanac, K.E., Pieterse, A.H., Price, A., Rabben, J., Riganti, P., Sanatani, M., Scheibler, F., Schoefs, E., Taylor, O.A., Valentine, K.D., Wexler, R., 2025. Shared Decision-Making. A Primer for Clinicians. J. Gen. Intern. Med. 40, 3889–3899. 10.1007/s11606-025-09707-z

Franco, J.V.A., Riganti, P., Yanzi, M.V.R., Kopitowski, K., 2020. Equipoise is preference sensitive. Can. Fam. physician Med. Fam. Can. 66, 551–552.

Harris, P.A., Taylor, R., Minor, B.L., Elliott, V., Fernandez, M., O’Neal, L., McLeod, L., Delacqua, G., Delacqua, F., Kirby, J., Duda, S.N., Consortium, on behalf of the Redc., 2019. The REDCap Consortium: Building an International Community of Software Platform Partners. J Biomed Inform 95, 103208. 10.1016/j.jbi.2019.103208

Harris, P.A., Taylor, R., Thielke, R., Payne, J., Gonzalez, N., Conde, J.G., 2009. Research electronic data capture (REDCap)—A metadata-driven methodology and workflow process for providing translational research informatics support. J Biomed Inform 42, 377–381. 10.1016/j.jbi.2008.08.010

Johansen, S.K., Kanstrup, A.M., Haseli, K., Stenmo, V.H., Thomsen, J.L., Rathleff, M.S., 2023. Exploring User Visions for Modeling mHealth Apps Toward Supporting Patient-Parent-Clinician Collaboration and Shared Decision-making When Treating Adolescent Knee Pain in General Practice: Workshop Study. Jmir Hum Factors 10, e44462. 10.2196/44462

Johansen, S.K., Maclachlan, L., Hillier, R., Taylor, G., Mellor, R., Rathleff, M.S., Vicenzino, B., 2022. Exploring patients’ and physiotherapists’ visions on modelling treatments and optimising self-management strategies for patellofemoral pain: A future workshop approach. Musculoskelet. Sci. Pr. 60, 102567. 10.1016/j.msksp.2022.102567

Joseph-Williams, N., Elwyn, G., Edwards, A., 2014. Knowledge is not power for patients: A systematic review and thematic synthesis of patient-reported barriers and facilitators to shared decision making. Patient Educ. Couns. 94, 291–309. 10.1016/j.pec.2013.10.031

Juel, N.G., Natvig, B., 2014. Shoulder diagnoses in secondary care, a one year cohort. Bmc Musculoskelet Di 15, 89. 10.1186/1471-2474-15-89

Jungk, R., Muellert, N.R., 1987. Future workshops: how to create desirable futures. London, England.

Khan, A., Husain, S.O., Kaur, R., Hashmat, A., Adil, A.N.K., Berkhamova, A., Awan, S.K., Varrassi, G., 2025. Primary Care Strategies for Managing Musculoskeletal Pain: A Narrative Overview. Cureus 17, e88447. 10.7759/cureus.88447

Larsen, J.B., Borregaard, P., Thomsen, J.L., Rathleff, M.S., Johansen, S.K., 2024. Improving general practice management of patients with chronic musculoskeletal pain: Interdisciplinarity, coherence, and concerns. Scand. J. Pain 24, 20230070. 10.1515/sjpain-2023-0070

Lucas, J., Doorn, P. van, Hegedus, E., Lewis, J., Windt, D. van der, 2022. A systematic review of the global prevalence and incidence of shoulder pain. BMC Musculoskelet. Disord. 23, 1073. 10.1186/s12891-022-05973-8

Lyng, K.D., Bengtsen, S.C., Zadro, J.R., Foster, N.E., Olesen, J.L., Thomsen, J.L., Søndergaard, J., Elwyn, G., Evans, R.E., Malliaras, P., Lowry, V., Desmeules, F., Rathleff, M.S., 2026. Development and initial evaluation of a patient decision aid to support decision-making in care-seeking patients with subacromial pain syndrome in primary care. Musculoskelet. Sci. Pract. 84, 103595. 10.1016/j.msksp.2026.103595

Lyng, K.D., Børsting, T.K., Clausen, M.B., Larsen, A.H., Liaghat, B., Ingwersen, K.G., Bateman, M., Rangan, A., Bjørnholdt, K.T., Christiansen, D.H., Jensen, S.L., Thomsen, J.L., Thorborg, K., Ziegler, C., Olesen, J.L., Rathleff, M.S., 2025. Shouldering our Way into a More Meaningful Research Agenda for Atraumatic Shoulder Pain: A Priority Setting Study. J. Orthop. Sports Phys. Ther. 1–36. 10.2519/jospt.2025.13059

Montori, V., Huijgens, F., Gionfriddo, M.R., Gravholt, D.L., Espinoza, N.R., Torres, R.L., Prokop, L., Ridgeway, J.L., Montori, V.M., Kunneman, M., 2026. The burden of shared decision-making: A scoping review of burden assessments in SDM research. Patient Educ. Couns. 147, 109550. 10.1016/j.pec.2026.109550

Montori, V.M., Ruissen, M.M., Hargraves, I.G., Brito, J.P., Kunneman, M., 2023. Shared decision-making as a method of care. BMJ Évid.-Based Med. 28, 213–217. 10.1136/bmjebm-2022-112068

(NICE), N.I. for H. and C.E., 2021. Shared decision-making. National Institute for Health and Care Excellence (NICE).

Powell, J.K., Lewis, J., Schram, B., Hing, W., 2024. Is exercise therapy the right treatment for rotator cufflrelated shoulder pain? Uncertainties, theory, and practice. Musculoskelet. Care 22, e1879. 10.1002/msc.1879

Powell, J.K., Schram, B., Lewis, J., Hing, W., 2022. “You have (rotator cuff related) shoulder pain, and to treat it, I recommend exercise.” A scoping review of the possible mechanisms underpinning exercise therapy. Musculoskelet. Sci. Pr. 62, 102646. 10.1016/j.msksp.2022.102646

RB, N., SK, J., J, A., TS, P., A, C., Rathleff, M.S., 2025. Integrating physiotherapists, occupational therapists, and psychologists in the management of chronic and complex musculoskeletal pain in the municipality – a workshop study. Musculoskelet. Sci. Pr. 78, 103336. 10.1016/j.msksp.2025.103336

Rhon, D.I., Horn, M.E., Lee, H.-J., Morton-Oswald, S., George, S.Z., 2025. Treatment variability for shoulder pain between physician and non-physician clinicians based on initial setting and specific shoulder diagnosis: a health system analysis. BMC Heal. Serv. Res. 25, 1370. 10.1186/s12913-025-13175-w

Skivington, K., Matthews, L., Simpson, S.A., Craig, P., Baird, J., Blazeby, J.M., Boyd, K.A., Craig, N., French, D.P., McIntosh, E., Petticrew, M., Rycroft-Malone, J., White, M., Moore, L., 2021. A new framework for developing and evaluating complex interventions: update of Medical Research Council guidance. Bmj 374, n2061. 10.1136/bmj.n2061

Stacey, D., Lewis, K.B., Smith, M., Carley, M., Volk, R., Douglas, E.E., Pacheco-Brousseau, L., Finderup, J., Gunderson, J., Barry, M.J., Bennett, C.L., Bravo, P., Steffensen, K., Gogovor, A., Graham, I.D., Kelly, S.E., Légaré, F., Sondergaard, H., Thomson, R., Trenaman, L., Trevena, L., 2024. Decision aids for people facing health treatment or screening decisions. Cochrane Database Syst. Rev. 2024, CD001431. 10.1002/14651858.cd001431.pub6

Storey, D., Collett, Castro-Lopes, J., Johnson, M., 2013. The challenges of pain management in primary care: a pan-European survey. J. Pain Res. 6, 393–401. 10.2147/jpr.s41883

Thomas, D.R., 2006. A General Inductive Approach for Analyzing Qualitative Evaluation Data. Am J Eval 27, 237–246. 10.1177/1098214005283748

Tong, A., Sainsbury, P., Craig, J., 2007. Consolidated criteria for reporting qualitative research (COREQ): a 32-item checklist for interviews and focus groups. Int. J. Qual. Heal. Care 19, 349–357. 10.1093/intqhc/mzm042

Vidal, R.V.V., 2005. The future workshopl: democratic problem solving, 1. oplag. ed. IMM, Informatik og Matematisk Modelling, DTU, Lyngby.

Windt, D.A. van der, Koes, B.W., Jong, B.A. de, Bouter, L.M., 1995. Shoulder disorders in general practice: incidence, patient characteristics, and management. Ann. Rheum. Dis. 54, 959. 10.1136/ard.54.12.959

